# Does the occurrence of a severe asthma exacerbation change the rate of subsequent events?

**DOI:** 10.1101/2021.04.05.21254930

**Authors:** Tae Yoon Lee, John Petkau, Mohsen Sadatsafavi

## Abstract

**Background:** Severe exacerbations requiring hospitalization are an important component of the natural history of asthma and a major source of its burden. Whether the occurrence of a severe exacerbation affects the rate of subsequent events has far-reaching implications in asthma management.

**Methods:** Using the centralized administrative health databases of British Columbia, Canada (1997/01/01 –2016/03/31), we created an incidence cohort of patients with at least one severe asthma exacerbation, defined as an episode of hospitalization with asthma as the primary diagnosis. We used an accelerated failure time joint frailty model for the time intervals between severe asthma exacerbations. Analyses were conducted separately for pediatric (< 14 years old) and adult (≥14 years old) patients.

**Results:** There were 3,039 patients (mean age at baseline 6.4, 35% female) in the pediatric group and 5,459 patients (mean age at baseline 50.8, 68% female) in the adult group, with 16% and 15%, respectively, experiencing at least one severe asthma exacerbation during follow-up. The first follow-up severe asthma exacerbation was associated with an increase of 79% (95% CI: 15% – 186%) in the rate of the subsequent events for the pediatric group. The corresponding value was 186% (95% CI: 85% – 355%) for the adult group. For both groups, the effects of subsequent severe exacerbations were not statistically significant.

**Conclusion:** Our findings suggest that among patients who have experienced their first severe asthma exacerbation, preventing the next event can drastically change the course of the disease and reduce the burden of future exacerbations.

## INTRODUCTION

Asthma is an early onset, life-long chronic disease of the airways responsible for a considerable impact on quality of life and a substantial economic burden[1]. Typical asthma symptoms include chest tightness, coughing, and shortness of breath[2]. Episodes of acute worsening of such symptoms, referred to as exacerbations or ‘flare-ups’, constitute an important component of the natural history of asthma[3]. In particular, severe exacerbations are one of the main causes of emergency department visits, hospital admissions, missed school days, and loss of work productivity[4]. Even though the rate of severe exacerbations has been declining over the past decade in many countries, severe exacerbations continue to be a leading cause of hospitalization[5, 6].

In addition to short-term morbidity, an exacerbation might cause long-term structural damage to the lung. For example, a study of patients with moderate/severe asthma showed that one additional severe exacerbation per year was significantly associated with a permanent decline of 30.2 mL in forced expiratory volume at one second[7]. An important question in this context is whether the occurrence of a severe exacerbation affects the rate of subsequent events. Answering this question will not only provide insight into the natural history of asthma, but will also help properly assess the value of preventive therapies. For instance, anti-inflammatory therapies are well-known to reduce the rate of asthma exacerbations[8]. If an exacerbation increases the rate of subsequent events, then the benefit of anti-inflammatory therapies will transcend the prevention of the ‘next’ exacerbation into future ones, even after the therapy is discontinued. This will affect the risk-benefit and cost-effectiveness of such therapies.

To the best of our knowledge, this important question has not been evaluated in asthma. Conventional wisdom is that previous history of asthma exacerbations is the best predictor of future exacerbations[3, 9]. However, in the presence of between-patient variability in exacerbation rates, the previous exacerbation history provides phenotypic information about a patient (e.g., whether the patient is a frequent exacerbator), which itself predicts the future exacerbation rate. Consequently, the strong correlation between past and future exacerbations, as documented by previous investigators, does not imply causation. In this paper, we report on a population-based retrospective cohort study to specifically determine the effect of severe asthma exacerbations on the rate of subsequent events, separately within paediatric and adult asthma patients.

## METHODS

### Data sources

We used the centralised administrative databases of British Columbia, a province of Canada with a population of 4.6 million as of 2016[10]. All the legal residents of the province are included in the databases, minimising the risk of selection bias, and the quality of the data is high with a very low level of missing, under-reported, or misclassified entries[11]. We had access to the following databases for the period of 1997/01/01 to 2016/03/31: demographics, including socio-economic status (SES) based on the neighborhood income quintile[12], registration status with the provincial healthcare system[12], vital statistics[13], and inpatient and outpatient records[14, 15]. All inferences, opinions, and conclusions drawn in this paper are those of the authors, and do not reflect the opinions or policies of the Data Steward(s). Ethics approval was obtained from the Human Ethics Board of the University of British Columbia (H17-00938).

### Study cohort

To construct an incidence cohort of asthma patients, we first identified all patients who experienced at least one severe asthma exacerbation. A severe asthma exacerbation was defined as an episode of hospitalization with asthma as the primary diagnosis using the International Classification for Disease codes (ICD, 9th revision 493, excluding 493.2x; 10th revision J45-46); we treated hospitalizations occurring within 7 days of each other as one severe exacerbation event. We chose this definition as the entry criterion due to its high positive predictive value. It also results in a relatively homogeneous severe asthma cohort, reducing the confounding by disease severity. We refer to the first severe asthma exacerbation as the index exacerbation and the corresponding date as the index date, marking the beginning of follow-up. Subsequent severe asthma exacerbations are referred to as follow-up events. To ensure that the first asthma-related hospitalization in the data was indeed the first instance of a severe exacerbation, we excluded patients who had less than five years of continuous presence in the study period before index date[5]. Similarly, we excluded patients younger than 5 years of age on index date if they were not continuously present in the registration database within this period. We also excluded patients with a history of hospitalization due to chronic obstructive pulmonary disease (COPD; ICD, 9th version 491, 492, 493.2x, 496; 10th version J41-44) before index date, as the diagnostic likelihood and natural history of asthma exacerbations in COPD patients might be different from that in the general asthma population[16]. Finally, the cohort was divided into the subgroups of paediatric and adult patients based on age on the index date, given the potentially different disease dynamics and associations in the two subgroups[17]. The paediatric group consisted of patients less than 14 years of age on index date, whereas the adult group included older patients. All patients were followed until the first occurrence of the following: the last date of any medical service use record, de-registration from the health care system, a hospital admission due to COPD, death, and the end of study period.

### Exposure, outcome, and covariates

The exposure was the occurrence of a severe asthma exacerbation requiring hospital admission, and the outcome was the time to the next severe asthma exacerbation. In addition to asthma exacerbations, we modeled hospitalization due to COPD and all-cause mortality as competing events for the adult group (these events were infrequent for the paediatric group and thus were not explicitly modelled; see Table 1), given their potential association with asthma[18, 19]. In our study, the competing events are terminal events that stop further observation of severe asthma exacerbations. During a severe asthma exacerbation episode, we considered a patient to be at risk for death but not at risk of another severe asthma exacerbation or a severe COPD exacerbation.

**Table 1.**
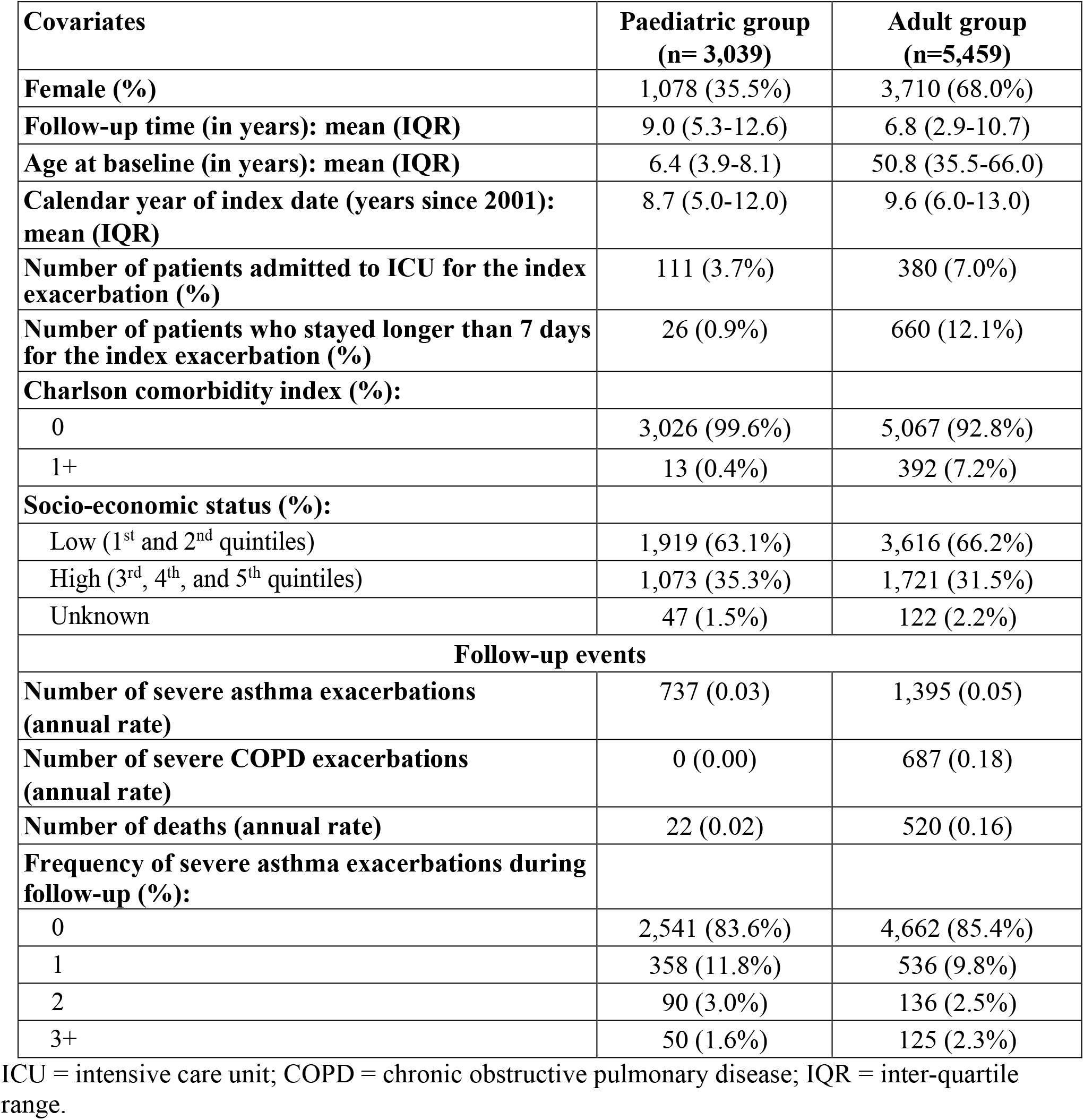
Summary statistics of the key covariates and outcomes of the final cohort.

| **Covariates** | **Paediatric group**<br>**(n= 3,039)** | **Adult group**<br>**(n=5,459)** |
| --- | --- | --- |
| **Female (%)** | 1,078 (35.5%) | 3,710 (68.0%) |
| **Follow-up time (in years): mean (IQR)** | 9.0 (5.3-12.6) | 6.8 (2.9-10.7) |
| **Age at baseline (in years): mean (IQR)** | 6.4 (3.9-8.1) | 50.8 (35.5-66.0) |
| **Calendar year of index date (years since 2001): mean (IQR)** | 8.7 (5.0-12.0) | 9.6 (6.0-13.0) |
| **Number of patients admitted to ICU for the index exacerbation (%)** | 111 (3.7%) | 380 (7.0%) |
| **Number of patients who stayed longer than 7 days for the index exacerbation (%)** | 26 (0.9%) | 660 (12.1%) |
| **Charlson comorbidity index (%):** |  |  |
| 0 | 3,026 (99.6%) | 5,067 (92.8%) |
| 1+ | 13 (0.4%) | 392 (7.2%) |
| **Socio-economic status (%):** |  |  |
| Low (1<sup>st</sup> and 2<sup>nd</sup> quintiles) | 1,919 (63.1%) | 3,616 (66.2%) |
| High (3<sup>rd</sup>, 4<sup>th</sup>, and 5<sup>th</sup> quintiles) | 1,073 (35.3%) | 1,721 (31.5%) |
| Unknown | 47 (1.5%) | 122 (2.2%) |
| **Follow-up events** |  |  |
| **Number of severe asthma exacerbations (annual rate)** | 737 (0.03) | 1,395 (0.05) |
| **Number of severe COPD exacerbations (annual rate)** | 0 (0.00) | 687 (0.18) |
| **Number of deaths (annual rate)** | 22 (0.02) | 520 (0.16) |
| **Frequency of severe asthma exacerbations during follow-up (%):** |  |  |
| 0 | 2,541 (83.6%) | 4,662 (85.4%) |
| 1 | 358 (11.8%) | 536 (9.8%) |
| 2 | 90 (3.0%) | 136 (2.5%) |
| 3+ | 50 (1.6%) | 125 (2.3%) |

ICU = intensive care unit; COPD = chronic obstructive pulmonary disease; IQR = inter-quartile range.

The analysis of association was controlled for the following covariates: index age, index calendar year, biological sex, index SES, and the Charlson comorbidity index calculated in the 12-month period before the index date[20]. The Charlson index was not included for the analysis in the paediatric group since only a handful had comorbidities (Table 1). For the primary event (severe asthma exacerbation), two additional binary covariates were included: whether a patient was admitted to the intensive care unit (ICU) for the index exacerbation, and whether the length of stay was 7 days or longer during the index exacerbation. These covariates were not included for COPD exacerbations and death due to the risk of immortal time bias[21].

### Statistical analysis

We used an accelerated failure time (AFT) joint frailty model for the time intervals or *gap times* between severe asthma exacerbations[22–24]. This framework is similar to that used previously to address the same question in the context of COPD[22]. The framework is particularly suitable for this analysis, as it accommodates the presence of heterogeneity in exacerbation rates across patients and the presence of competing risks, both of which are critical to control for to answer the question of interest[25, 26]. In this framework, the hazard function of severe asthma exacerbations is linked with the hazard functions of both competing events through a common, unobserved patient-specific random-effect. The random-effect represents the patient-specific propensity to exacerbate over and above the variability due to their observed covariates. A main advantage of using this framework is that such random-effects provide a mechanism to adjust for the potential confounding due to heterogeneity in exacerbation rates as well as unobserved covariates (e.g. symptom burden), thus enabling within-patient inference on the association between consecutive asthma exacerbations[27]. In an AFT model, the regression coefficient *β* for a covariate in the hazard function of the event of interest can be interpreted in terms of its rate acceleration factor (RAF), exp(*β*)[28]. For example, a RAF of 1.5 for a covariate means that each one unit increase in the covariate is associated with an increase in the rate of events of 50%. Details on our AFT joint frailty model can be found in Supplementary Material – Section 1.

AFT models require parametric specification of the baseline hazard function. We considered the generalised gamma baseline hazard function; popular hazard functions such as the exponential and Weibull are special cases[29]. We evaluated the goodness-of-fit using Akaike’s information criterion and visual comparison between the model-based and nonparametrically estimated cumulative incidence curves[30]. The model-based curve was obtained by taking the average of the predicted curves over all the patients. Details, including the final model fits, are provided in Supplementary Material - Section 2.

To evaluate the robustness of results against an alternative formulation for the competing events, we ran a sensitivity analysis for the adult group, whose details are provided in Supplementary Material – Section 3. Data analysis was performed using SAS, Version 9.4, and R, Version 3.6.1[31]. See Supplementary Material – Section 4 for details on the implementation of our AFT joint frailty model.

## RESULTS

### Patient characteristics

In the period from 1997/01/01 to 2016/03/13, there were 9,414 patients who experienced at least one severe asthma exacerbation after at least 5 years of continuous presence in the data (or with continuous presence in the data if they were younger than 5 years of age on index date). We excluded 887 patients who had a history of hospitalization due to COPD before index date, and 30 patients who had missing covariates. There were 3,039 patients in the paediatric group (index age < 14) and 5,459 patients in the adult group (index age ≥ 14) for the analysis. Figure 1 provides the schematic illustration of cohort creation.

**Figure 1:**
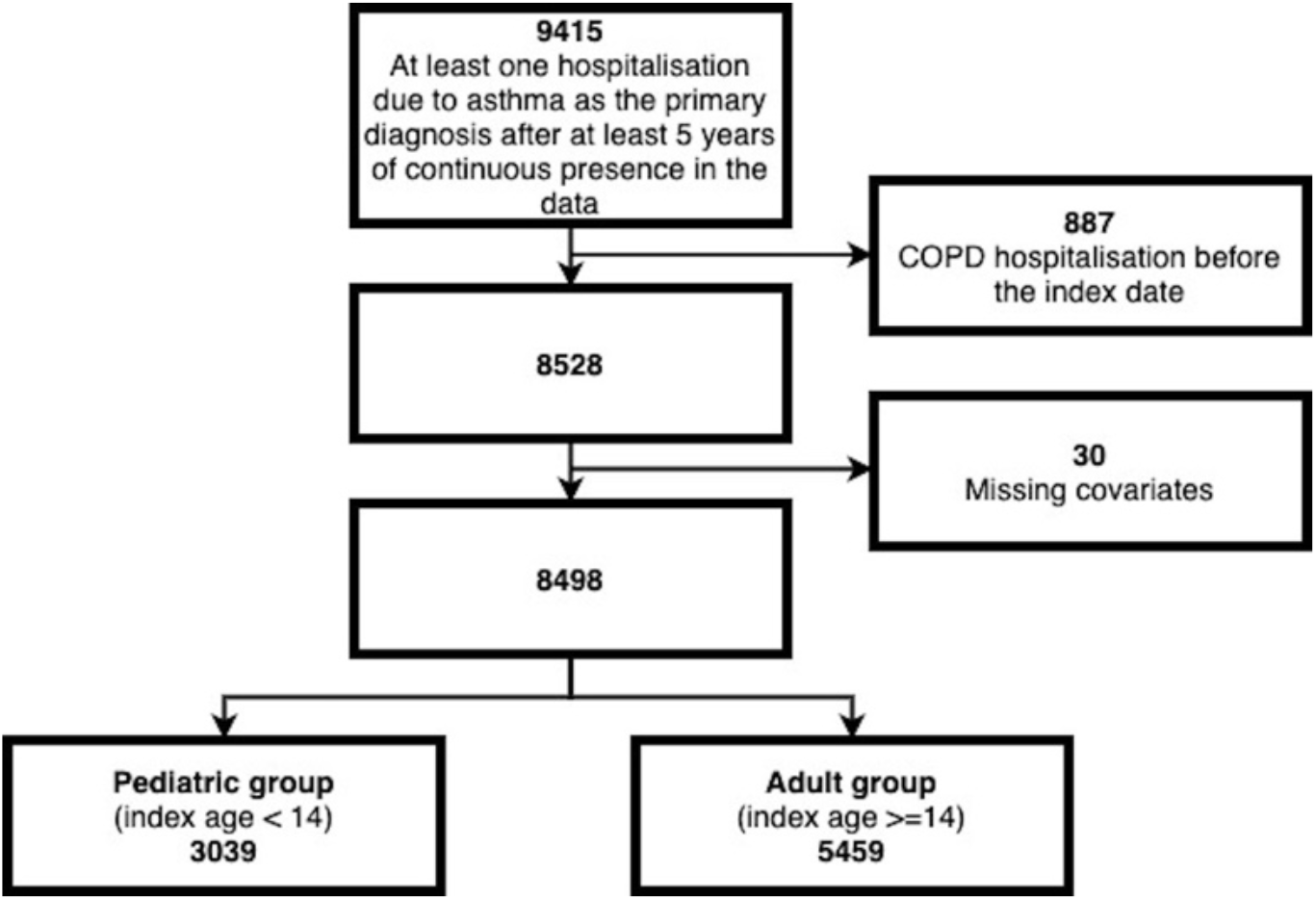
Flowchart of asthma cohort creation. COPD = chronic obstructive pulmonary disease

For the paediatric group, the mean index age was 6.4 years (inter-quartile range (IQR): 3.9-8.1 years), 35% were female, and the mean follow-up was 9.0 years (IQR: 5.3-12.6 years). A total of 737 follow-up severe asthma exacerbations were recorded, corresponding to an annualised rate of 0.03. 16% had at least one severe asthma exacerbation, 5% had two or more severe asthma exacerbations, and 2% had 3 or more severe exacerbations after index date. No severe COPD exacerbation was observed in this age group, and only 22 patients died during follow-up.

For the adult group, the mean index age was 50.8 years (IQR: 35.5-66.0 years), 68% were female, and the mean follow-up was 6.8 years (IQR: 2.9-10.7 years). A total of 1,395 follow-up severe asthma exacerbations occurred, corresponding to an annualised rate of 0.05. 15% had at least one severe asthma exacerbation, 5% had two or more severe asthma exacerbations, and 2% had 3 or more severe exacerbations after index date. During follow-up, 687 patients experienced severe COPD exacerbations, and 520 patients died. Table 1 presents the characteristics of both groups.

Among the models examined, those with the generalised gamma baseline hazard functions with gamma random-effects had the best goodness-of-fit in both the paediatric and adult groups for modeling the gap times between severe asthma exacerbations (Supplementary Material – Section 2).

Figure 2 provides the estimated rate acceleration factors (RAF) for the first three follow-up severe asthma exacerbations. For both groups, the occurrence of the first follow-up severe asthma exacerbation was associated with a significant increase in the rate of future events. For the paediatric group, the relative change in the rate was 79% (95% CI: 15-186%). This value was 186% (95% CI: 85 – 355%) for the adult group. The effects of subsequent severe exacerbations were not statistically significant for either group.

**Figure 2:**
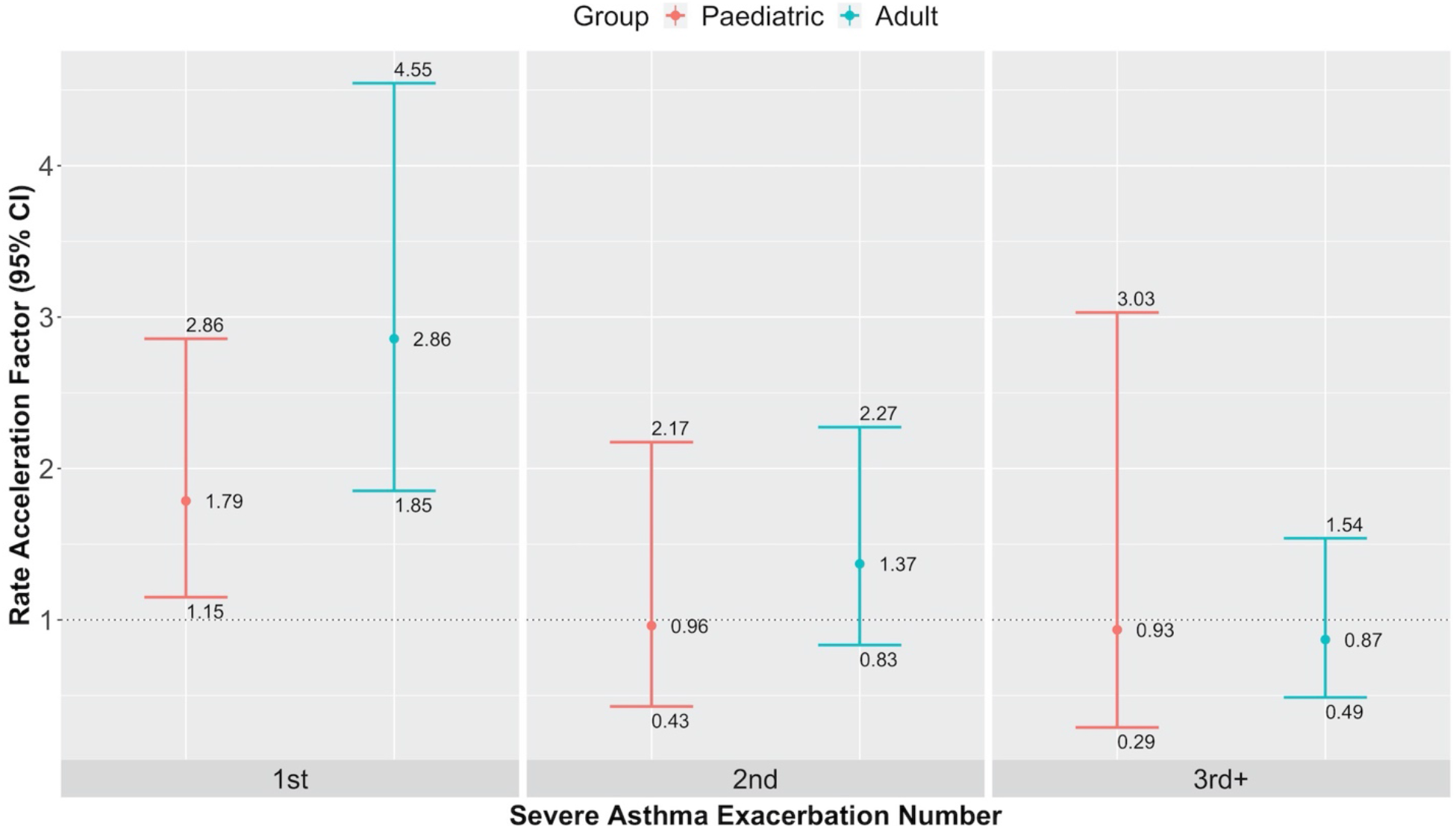
Estimated rate acceleration factors for severe asthma exacerbations in the paediatric (orange) and adult (blue) group. CI = confidence interval

Estimated model parameters and regression coefficients for all covariates are provided in Supplementary Material - Section 2. The sensitivity analysis demonstrated that results were not overly sensitive to the choices of time scale and hazard function used for the competing events in the adult group (see Supplementary Material – Section 3).

## DISCUSSION

Using a population-based cohort, we investigated the association between the occurrence of severe asthma exacerbations and the rate of subsequent severe asthma exacerbations, separately in the paediatric and adult groups. We found that the occurrence of the first follow-up severe exacerbation was associated with an increase in the rate of subsequent exacerbations by 79% and 186% in the paediatric and adult groups, respectively. The occurrences of the subsequent severe asthma exacerbations were not significantly associated with change in the exacerbation rate in either group, albeit there was a large degree of uncertainty around these estimates. Overall, our findings suggest that among patients who have had their first severe asthma exacerbation, preventing the next severe asthma exacerbation can significantly modify the trajectory of the disease and the rate of subsequent events, particularly in the adult group.

To the best of our knowledge, this is the first study on the associations between occurrences of severe asthma exacerbations and rate of subsequent events. Previous studies have evaluated the clustering of asthma exacerbations within individuals[3, 9, 32, 33]. For example, in a three-year prospective cohort study, the number of asthma exacerbations in the previous year was the strongest predictor of future asthma exacerbations[3]. However, such a finding does not indicate that the occurrence of an exacerbation modifies the rate (or risk) of future events. This is because in the presence of heterogeneity in exacerbation rates, a positive exacerbation history indicates that a patient is likely a ‘frequent exacerbator’, and hence is likely to exacerbate in the future. In other words, previous and future exacerbation rates are correlated, but correlation does not necessarily imply causation. In examining the effect of exacerbations on the rate of future events, the underlying rate of asthma exacerbation is a confounding factor that needs to be adjusted for. As described previously[27], the statistical model employed in this work allows for a patient-specific background exacerbation rate, thereby removing this confounding effect. This methodology also adjusts for any time-fixed variable that might associate previous and future asthma exacerbations.

Exacerbations can adversely modify the course of asthma through multiple mechanisms[34]. Neutrophils are prominent in the lungs during asthma exacerbations[35]. It is likely that proteolytic tissue destruction caused by neutrophil proteases will permanently scar the airways. Furthermore, epithelial damage is heightened during exacerbations[36]. It is postulated that the re-epithelialisation process can cause fibrosis through stimulating mesenchymal cells, increasing susceptibility to future exacerbations[34]. These mechanisms can result in narrowing of the airways and corresponding lung function decline, which is known to occur after severe asthma exacerbations[37]. However, the associations reported in this study are generally larger than one would expect from the effect of exacerbations on lung function alone. Beyond the effect on airways, other mechanisms might also be at play. For example, exacerbations are found to be associated with an acute loss of lung elasticity, which might not fully recover after the exacerbation is resolved[34]. This signals potentially structural damage to the lung and failure of tissue repair after an exacerbation.

This study has several strengths. First, we studied a population-based incidence cohort, which is generally free of selection bias. Second, we defined cohort entry based on the first instance of asthma-related hospitalization to obtain a homogenous cohort, reducing the confounding effect of background severity of asthma. Previous chart review studies have shown that this definition has a high positive predictive value for identifying asthma patients[38, 39], minimising the risk of exposure and outcome misclassification. Third, we used a robust statistical methodology to allow for variation beyond and over that explained by covariates through random-effects. Moreover, the methodology enabled us to cope with competing events of severe COPD exacerbations and death in the adult group, avoiding biases from treating competing events as non-informative censoring[26].

This study also has limitations. Despite the relatively large cohort, we had a modest number of severe asthma exacerbations after the first follow-up event. Consequently, we could not provide precise estimates on the change in the exacerbation rate due to subsequent exacerbations. In a similar study for COPD, with more than 34,000 follow-up severe COPD exacerbations, Sadatsafavi et al showed a positive, diminishing effect of occurrences of successive follow-up severe COPD exacerbations on the rate of subsequent events, which remained significant up to the fourth follow-up event[22]. To further refine the current results, future research utilising a larger dataset is required. To increase the number of events, we could have included exacerbations that were managed in emergency departments or at physician clinics (e.g., with the use of systemic corticosteroids). However, verifying such events as asthma exacerbations is not easy in our databases. One concern was that patients might receive systemic corticosteroids for other conditions (prescription records are not accompanied by a reason for the prescription). Moreover, a discharge code is not provided for an emergency department visit, making it difficult to identify the underlying reason for the visit. Next, we used the index exacerbation as an entry criterion into the cohort. While this entry criterion provided a cohort definition with high specificity, it prevented us from evaluating the effect of the index exacerbation on subsequent outcomes. Furthermore, we did not adjust our analyses for treatment because of the difficulty with incorporating its time-varying effect in AFT frailty models. If the occurrence of an exacerbation prompted the initiation of anti-inflammatory therapies, the rate of a subsequent event would most likely decrease. Hence, not adjusting for treatment should move the estimated RAF toward the null. In consideration of this, our finding that the first follow-up severe asthma exacerbation increases the rate of subsequent events remains valid.

In conclusion, this study showed that severe exacerbations early in the course of asthma are associated with an increase in the rate of subsequent severe exacerbations, in both children and adults. Besides offering a new insight into the natural history of asthma, these results have important implications for quantifying the benefit of preventive therapies. One implication of our findings is that if a preventive controller therapy (e.g., biologic indicated for severe asthma) is used for a certain period of time, its effect transcends beyond this period if it has prevented the occurrence of a severe exacerbation. This is critical when extrapolating the results from clinical trials (with relatively short follow-up time) towards quantifying the risk-benefit and cost-effectiveness of asthma therapies.

## Data Availability

The data used in this study are from Population Data BC. They are available to the public under restrictions, according to British Columbia's Freedom of Information and Protection of Privacy Acts.

https://www.popdata.bc.ca/privacy/policies/legislativeframework

## Acknowledgment

This study was funded by Genome Canada and Genome British Columbia - 274CHI. The funders did not have any role in any aspects of this study.

## Notes

### Competing Interest Statement

T.Y.L reports no conflict of interest. J.P. reports personal fees from Biogen, personal fees from Teva Pharmaceuticals Europe, personal fees from The Canadian Study Group on CCSVI, personal fees from Novartis, grants from Natural Sciences and Engineering Research Council of Canada (NSERC), outside the submitted work. M.S. has received honoraria from GlaxoSmithKline, Boehringer Ingelheim, Teva, and AstraZeneca. He has received research fund directly into his accounts at The University of British Columbia from AtraZeneca, Boehringer Ingelheim, and MethaPharm Inc.

### Author Declarations

Ethics approval was obtained from the Human Ethics Board of the University of British Columbia (H17-00938).

## REFERENCES

1. Ehteshami-Afshar S, FitzGerald JM, Doyle-Waters MM, Sadatsafavi M. The global economic burden of asthma and chronic obstructive pulmonary disease. Int. J. Tuberc. Lung Dis. 2016; 20: 11–23.

2. Pearce N, Aït-Khaled N, Beasley R, Mallol J, Keil U, Mitchell E, Robertson C. Worldwide trends in the prevalence of asthma symptoms: phase III of the International Study of Asthma and Allergies in Childhood (ISAAC). Thorax BMJ Publishing Group Ltd; 2007; 62: 758–766.

3. Chipps BE, Zeiger RS, Borish L, Wenzel SE, Yegin A, Hayden ML, Miller DP, Bleecker ER, Simons FER, Szefler SJ, others. Key findings and clinical implications from The Epidemiology and Natural History of Asthma: Outcomes and Treatment Regimens (TENOR) study. Journal of Allergy and Clinical Immunology Elsevier; 2012; 130: 332– 342.

4. Bahadori K, Doyle-Waters MM, Marra C, Lynd L, Alasaly K, Swiston J, FitzGerald JM. Economic burden of asthma: a systematic review. BMC Pulmonary Medicine Springer; 2009; 9: 24.

5. Public Agency of Canada. Report from the Canadian chronic disease surveillance system: asthma and chronic obstructive pulmonary disease in Canada [Internet]. 2018 [cited 2020 Mar 9].Available from: http://publications.gc.ca/collections/collection_2018/aspc-phac/HP35-90-2018-eng.pdf.

6. Global Asthma Network. The global asthma report 2018. [Internet]. 2018 [cited 2020 Jun 30].Available from: http://www.globalasthmareport.org/Global%20Asthma%20Report%202018.pdf.

7. Bai T, Vonk J, Postma D, Boezen H. Severe exacerbations predict excess lung function decline in asthma. European Respiratory Journal Eur Respiratory Soc; 2007; 30: 452–456.

8. Horvath G, Wanner A. Inhaled corticosteroids: effects on the airway vasculature in bronchial asthma. European Respiratory Journal Eur Respiratory Soc; 2006; 27: 172–187.

9. Schatz M, Meckley LM, Kim M, Stockwell BT, Castro M. Asthma exacerbation rates in adults are unchanged over a 5-year period despite high-intensity therapy. Journal of Allergy and Clinical Immunology: In Practice Elsevier; 2014; 2: 570–574.

10. Statistics Canada. British Columbia [Province] and Canada [Country] (table). Census profile. 2016 Census. Statistics Canada Catalogue no. 98-316-X2016001. Ottawa. Statistics Canada; 2017.

11. Canadian Institute for Health Information. Data quality study of the 2015–2016 discharge abstract database: a focus on hospital harm. Ottawa, ON: CIHI Canadian Institute for Health Information; 2016.

12. British Columbia Ministry of Health [creator] (2017). Consolidation file (MSP registration & premium billing). V2. Population Data BC [publisher]. Data Extract. MOH. [Internet]. 2017.Available from: http://www.popdata.bc.ca/data.

13. BC Vital Statistics Agency [creator] (2017). Vital statistics deaths. V2. Population Data BC [publisher]. Data Extract. BC Vital Statistics Agency [Internet]. 2017.Available from: http://www.popdata.bc.ca/data.

14. British Columbia Ministry of Health [creator] (2017). Medical services plan (MSP) payment information file. V2. Population Data BC [publisher]. Data Extract. MOH. [Internet]. 2017.Available from: http://www.popdata.bc.ca/data.

15. Canadian Institute for Health Information [creator] (2017). Discharge abstract database (Hospital Separations). V2. Population Data BC [publisher]. Data Extract. MOH. [Internet]. 2017.Available from: http://www.popdata.bc.ca/data.

16. Tho NV, Park HY, Nakano Y. Asthma–COPD overlap syndrome (ACOS): a diagnostic challenge. Respirology Wiley Online Library; 2016; 21: 410–418.

17. Trivedi M, Denton E. Asthma in children and adults–what are the differences and what can they tell us about asthma? Frontiers in Pediatrics Frontiers; 2019; 7: 256.

18. Silva GE, Sherrill DL, Guerra S, Barbee RA. Asthma as a risk factor for COPD in a longitudinal study. Chest Elsevier; 2004; 126: 59–65.

19. Diaz-Guzman E, Khosravi M, Mannino DM. Asthma, chronic obstructive pulmonary disease, and mortality in the US population. COPD: Journal of Chronic Obstructive Pulmonary Disease Taylor & Francis; 2011; 8: 400–407.

20. Romano PS, Roos LL, Jollis JG. Presentation adapting a clinical comorbidity index for use with ICD-9-CM administrative data: differing perspectives. Journal of Clinical Epidemiology Elsevier; 1993; 46: 1075–1079.

21. Suissa S. Immortal time bias in pharmacoepidemiology. American Journal of Epidemiology Oxford University Press; 2008; 167: 492–499.

22. Sadatsafavi M, Xie H, Etminan M, Johnson K, FitzGerald JM, Canadian Respiratory Research Network. The association between previous and future severe exacerbations of chronic obstructive pulmonary disease: updating the literature using robust statistical methodology. PloS One 2018; 13: e0191243.

23. Liu L, Wolfe RA, Huang X. Shared frailty models for recurrent events and a terminal event. Biometrics 2004; 60: 747–756.

24. Huang X, Liu L. A joint frailty model for survival and gap times between recurrent events. Biometrics 2007; 63: 389–397.

25. Sadatsafavi M, FitzGerald JM. Heterogeneity’s ruses: the neglected role of between-individual variability in longitudinal studies of COPD exacerbations. Thorax 2014; 69: 1043–1044.

26. Lau B, Cole SR, Gange SJ. Competing risk regression models for epidemiologic data. American Journal of Epidemiology Oxford University Press; 2009; 170: 244–256.

27. Duchateau L, Janssen P. The frailty model. Springer Science & Business Media; 2007.

28. Wei L-J. The accelerated failure time model: a useful alternative to the Cox regression model in survival analysis. Statistics in Medicine Wiley Online Library; 1992; 11: 1871–1879.

29. Cox C, Chu H, Schneider MF, Munoz A. Parametric survival analysis and taxonomy of hazard functions for the generalized gamma distribution. Statistics in Medicine Wiley Online Library; 2007; 26: 4352–4374.

30. Benichou J, Gail MH. Estimates of absolute cause-specific risk in cohort studies. Biometrics JSTOR; 1990; : 813–826.

31. R Core Team. R: a language and environment for statistical computing. Vienna, Austria; 2019.

32. Gershon A, Guan J, Victor JC, Wang C, To T. The course of asthma activity: a population study. Journal of Allergy and Clinical Immunology Elsevier; 2012; 129: 679–686.

33. Schatz M, Zeiger RS, Mosen D, Vollmer WM. Asthma-specific quality of life and subsequent asthma emergency hospital care. American Journal of Managed Care 2008; 14: 206–211.

34. Rennard SI, Farmer SG. Exacerbations and progression of disease in asthma and chronic obstructive pulmonary disease. Proc Am Thorac Soc 2004; 1: 88–92.

35. Fahy JV, Kim KW, Liu J, Boushey HA. Prominent neutrophilic inflammation in sputum from subjects with asthma exacerbation. J. Allergy Clin. Immunol. 1995; 95: 843–852.

36. Picado C. Classification of severe asthma exacerbations: a proposal. Eur. Respir. J. 1996; 9: 1775–1778.

37. Matsunaga K, Hirano T, Oka A, Tanaka A, Kanai K, Kikuchi T, Hayata A, Akamatsu H, Akamatsu K, Koh Y, Nakanishi M, Minakata Y, Yamamoto N. Progression of Irreversible Airflow Limitation in Asthma: Correlation with Severe Exacerbations. J Allergy Clin Immunol Pract 2015; 3: 759-764.e1.

38. Firoozi F, Lemière C, Beauchesne M, Forget A, Blais L. Development and validation of database indexes of asthma severity and control. Thorax 2007; 62: 581–587.

39. Prosser RJ, Carleton BC, Smith MA. Identifying persons with treated asthma using administrative data via latent class modelling. Health Serv Res 2008; 43: 733–754.

