## Supplementary Materials for "Does the occurrence of a severe asthma exacerbation change the rate of subsequent events?"

### 1 Details of statistical analysis

Severe asthma exacerbations are recurring events. In our study design, the follow-up of the asthma exacerbations can be stopped by two main terminal events: a severe COPD exacerbation (as an indicator that fixed airflow obstruction has occurred) and death. Studies show that asthma exacerbations and each of the terminal events are not independent; asthma exacerbations increase the risk of COPD and death[1, 2]. Consequently, treating these competing risks as non-informative censoring could lead to potential biases in analysis of the recurrent event process of severe asthma exacerbations[3].

To achieve our objectives in a unified framework, we used a joint frailty model that is specifically designed to deal with recurrent events and competing risks[4]. In simple terms, the hazard function for severe asthma exacerbations was linked with the hazard function for death and for severe COPD exacerbation via a common patient-specific random-effect. The random-effects allow for the heterogeneity in the severe asthma exacerbation rates and characterise the dependence between the recurrent event process and each of the terminal event processes. We used accelerated failure time (AFT) type hazard functions for more intuitive interpretability of the regression coefficients in the hazard functions[5].

#### 1.1 Definitions and notation

For each patient $i = 1, \dots, n$, let $T_{i,j}$ be observed event times since the starting date of their index exacerbation (referred to as the *index date*) for $j = 1, \dots, m_i + 1$, where $m_i$ is the (random) number of recurrent events that patient $i$ experiences. Note that the last event cannot be a severe asthma exacerbation since the follow-up of severe asthma exacerbations is stopped by either a severe COPD exacerbation, death, or the end of the study period. To fix the follow-up time scale, we set the index date as 0 for all patients, i.e. $T_{i,0} = 0$.

Let $C_i$, $D_i$, and $CR_i$ be the time since the index date of the start of the first severe COPD exacerbation, death, and censoring, respectively, for patient $i$. Let $FU_i = \min(C_i, D_i, CR_i)$ be the patient's follow-up time. While the exact date was available for all the events, date of birth and death were only available by the year and month due to privacy reasons. For birth date, we used the 15th of the month as a default. As for the death date, we used the day of the last medical resource use in the death month; if no medical resource was used, then we used the last day of the death month as a default.

For each patient, a 'gap time' was defined as the time from the beginning (the admission date) of a severe asthma exacerbation to the beginning (the admission date) of the next severe asthma exacerbation, the admission date of a severe COPD exacerbation, the date of death, or the end of follow-up, whichever occurred first. We formally define gap times between consecutive event times by $W_{i,j} = T_{i,j} - T_{i,j-1}$ for $j = 1, \dots, m_i + 1$, where $T_{i,0} = 0$ and $T_{i,m_i+1} = FU_i$. Note that the gap times were defined by occurrence of exacerbations (except for the last observed event that terminated further observation of severe asthma exacerbations). We used the gap times as the scale of measurement as the purpose of the study is to examine the effects of both time-dependent and time-independent covariates on the times to subsequent events. For the competing events, the gap times represent the time intervals of the calendar times decomposed by the occurrence of severe asthma exacerbations. In other words, we used *residual calendar (time-to-event) times* for the competing events.

The $j$th gap time $W_{i,j}$ contains the duration of the $(j-1)$th severe asthma exacerbation, $B_{i,j-1}$. We assume that patients are not at risk for another severe asthma exacerbation or for a severe COPD exacerbation while experiencing a severe asthma exacerbation. To reflect this, we used modified gap times with these durations excluded for severe asthma and COPD exacerbations: $W_{i,j}^{*} = W_{i,j} - B_{i,j-1}$. On the other hand, we assume that patients are still at risk for death while experiencing a severe asthma exacerbation. Let $\delta_i^C$ and $\delta_i^D$ be the indicator functions that are equal to 1 when the last observed event that patient $i$ experiences is a severe COPD exacerbation and death, respectively.

We assume that 1) the recurrent event process and the two terminal event processes are independent given the (unobserved) random-effect and covariates and that censoring (due to other reasons) is independent of those processes. This framework further assumes that that 2) the modified inter-exacerbation times, $W_{i,j}^{*}$, of asthma are independent and identically distributed (i.i.d.) given the covariates and the random-effect and 3) the residual calendar times to the first severe COPD exacerbation and death (defined by the recurrent event process) possess the *memoryless property*. While the first two assumptions are common in the literature, the third assumption on the residual times is specific to our formulation which uses the gap times as the scale of measurement for the terminal processes as well as the recurrent event process.

To elaborate on the memoryless property of the residual calendar times, consider the terminal event process of death for patient $i$. For any $t > 0$ and $s > 0$, a random variable $D_i$ has the memoryless property if $P(D_i > s + t \mid D_i \geq t) = P(D_i > s)$. The memoryless property of the residual calendar times for the death process, together with the independence of the recurrent event and death processes, implies that for the $j$th event time $T_{i,j}$ ($j = 1, \dots, m_i$) and any $s > 0$, $P(D_i - T_{i,j} > s \mid D_i \geq T_{i,j}) = P(D_i > s + T_{i,j} \mid D_i \geq T_{i,j}) = P(D_i > s)$. This implies that the hazard for death 'resets' after each recurrent event but also requires that $D_i$ follow the exponential distribution, the only (continuous) distribution having the memoryless property[6]. To further validate the estimated regression coefficients for the adult group (for which the competing events are modeled), we ran a sensitivity analysis using calendar times, rather than residual times, for the competing events (Section 3).

#### 1.2 Joint frailty model

Denote the hazard function for the recurrent event process by $h(\cdot)$, for the terminal event process of severe COPD exacerbations by $h^C(\cdot)$, and for the terminal event process of death by $h^D(\cdot)$. Let $\epsilon_i$ ($i = 1, \dots, n$) be i.i.d. unobserved, patient-specific random-effects that allow for the heterogeneity in severe asthma exacerbation rates. The AFT-type joint frailty model adapted from [4] is

$$\begin{cases} h(w_{i,j}^*|\epsilon_i) = h_0\left(w_{i,j}^*\, \epsilon_i \exp\left(\beta_A^T Z_{i,j}^A\right)\right) \epsilon_i \exp\left(\beta_A^T Z_{i,j}^A\right) & \textbf{\textit{asthma}} \\ h^C(w_{i,j}^*|\epsilon_i) = h_0^C\left(w_{i,j}^*\, \epsilon_i^{\alpha_C} \exp(\beta_C^T Z_{i,j}^C)\right) \epsilon_i^{\alpha_C} \exp\left(\beta_C^T Z_{i,j}^C\right) & \textbf{\textit{COPD}} \\ h^D(w_{i,j}|\epsilon_i) = h_0^D\left(w_{i,j} \epsilon_i^{\alpha_D} \exp(\beta_D^T Z_{i,j}^D)\right) \epsilon_i^{\alpha_D} \exp(\beta_D^T Z_{i,j}^D) & \textbf{\textit{death}} \end{cases}$$

where $h_0$, $h_0^C$, and $h_0^D$ are the baseline hazard functions and $\beta_k$ are regression coefficients for the corresponding covariates $Z_{i,j}^k$ for $k = A$ (asthma), $C$ (COPD), and $D$ (death). The random-effect $\epsilon_i$ not only allows for the patient-specific heterogeneity, it also describes the dependence between the recurrent event process and the terminal event process of type $k$ through the exponents $\alpha_k$ that specify the strength and direction of the dependence for $k = C$ (COPD) and $D$ (death). We emphasise that is the only way the recurrent events are linked with the terminal events in our model. We require that $E(\epsilon_i) = 1$ so that the intercept parameter is identifiable in each sub-model and assume that $\text{Var}(\epsilon_i) = \theta > 0$.

AFT models offer an intuitive interpretation of regression coefficients. We define the accelerated failure time (AFT) factor of the regression coefficient $\beta_j$ as $\exp(-\beta_j)$. If the corresponding covariate is increased by one unit while all the other covariates remain constant, then the mean time to the events of interest is multiplied by the AFT factor. We can give an alternative interpretation in terms of the rate of the events of interest. We refer to the inverse of the AFT factor as the rate acceleration factor (RAF): RAF = $\exp(\beta_j)$. Then the percent relative change in the rate of subsequent events is given by [RAF-1]$*$ 100%.

#### 1.3 Likelihood

The hazard function $h(\cdot)$ determines three additional functions of importance: the cumulative hazard function, $H(w) = \int_0^w h(u)du$ , the survival function, $S(w) = \exp(-H(w))$, and the density function, $f(w) = S(w)h(w)$. Furthermore, let $\Phi = \{\beta_A, \beta_C, \beta_D, \alpha_C, \alpha_D\}$ and $\theta$ be the set of parameters we wish to estimate. For the remainder of the section, the density, hazard, and survival functions are conditional on the random-effect unless stated otherwise. We first derive the conditional likelihood for patient $i$ given the random-effect $\epsilon_i$. By definition, there are $m_i$ recurrent events. For each occurrence of the recurrent event process, the patient does not experience the two terminal events, a severe COPD exacerbation and death. Thus, the contribution to the likelihood corresponding to experiencing the $j$th recurrent event is the product of $f_i\left(w^*_{i,j}\right)$, $S_i^C\left(w^*_{i,j}\right)$, and $S_i^D\left(w_{i,j}\right)$. For the last $(m_i + 1)$th event, the patient either experiences a severe COPD exacerbation, dies, or exits the study (censored) without experiencing another severe asthma exacerbation. Thus, the conditional likelihood for patient $i$ is

$$L_i(\Phi|\epsilon_i) = [\ \Pi_{j=1}^{m_i} f_i\left(w^*_{i,j}\right) S_i^C\left(w^*_{i,j}\right) S_i^D\left(w_{i,j}\right)]$$
$$\times [\ S_i\left(w^*_{i,m_i+1}\right)$$

$$\times \left(f_i^C\left(w_{i,m_i+1}^*\right)\right)^{\delta_i^C} \left(S_i^C\left(w_{i,m+1}^*\right)\right)^{1-\delta_i^C}$$

$$\times \left(f_i^D\left(w_{i,m_i+1}\right)\right)^{\delta_i^D} \left(S_i^D\left(w_{i,m_i+1}\right)\right)^{1-\delta_i^D} ].$$

The marginal likelihood for patient $i$ is given by integrating out the random-effect $\epsilon_i \sim f_\theta(\cdot)$:

$$L_i(\Phi, \theta) = \int L_i(\Phi|\epsilon_i) f_\theta(\epsilon_i) d\epsilon_i.$$

Finally, assuming that patients are independent, the marginal likelihood for the whole sample is simply the product of the individual marginal likelihoods:

$$L(\Phi, \theta) = \Pi_{i=1}^{n} \mathrm{L_i}(\Phi, \theta).$$

#### 1.4 Estimation

We used SAS PROC NLMIXED[7] to obtain the maximum likelihood estimate of the parameters based on Gaussian quadrature techniques[8–10]. For the optimization method, we either used the conjugate gradient algorithm or the Newton-Raphson algorithm. We used two commonly used distributions for $\epsilon_i$: log-normal ($\mu = 0, \sigma^2 = \theta$) and gamma (shape=$\theta$, rate=$\theta$). As SAS PROC NLMIXED allows only normal random-effects, we made use of the probability integral transformation[11] to implement the gamma random-effect[12]. See the SAS code for implementation and more details in Section 4.

### 2 Model selection, goodness-of-fit, and final model fits

#### 2.1 Paediatric group

For the paediatric group (n=3,039), 16% had at least one severe asthma exacerbation, 5% had two or more severe asthma exacerbations, and 2% had 3 or more severe exacerbations after their index date. About 70% of the 737 follow-up severe asthma exacerbations were first exacerbations. Only the recurrent event process of severe asthma exacerbations was considered since no severe COPD exacerbation and a small number of deaths were observed.

We had to choose the form of the baseline hazard and type of random-effects. For the baseline hazard function, we considered the generalised gamma (GG) distribution, special cases of which include the log-normal, gamma and Weibull distributions[13]. For the random-effects, we considered gamma (G) and log-normal (LN) distributions as specified in Section 1.4. The GG models with G random-effects (GG-G) and LN random-effects (GG-LN) were fitted successfully. We assessed the goodness-of-fit of the models and compared the models using cumulative incidence curves (CICs[14]). In the absence of competing risks, as is the case here, the nonparametric estimate (ignoring covariates) of the CIC is equal to 1-KM(t) (Kaplan-Meier estimate). The model-based cumulative incidence curve and its 95% CI were obtained as described in Chapter 9 of [14]. From the model estimates, we first generated 100 samples of the random-effects for each patient based on the posterior distribution obtained from the empirical Bayes procedure in PROC NLMIXED, calculated the 100 CICs for each patient, took the average at each fixed time to obtain the patient-specific model-based CIC, and then finally took the average of the patient-specific model-based CICs at each fixed time to obtain the model-based CIC. Note that we integrated out the random-effects by the Monte-Carlo method (with 100 replications). For perfect goodness-of-fit, we would expect the nonparametric (NP) and model-based CICs to be incident upon each other. We used diagnostic CIC-based plots in preference to the Akaike information criterion (AIC) to compare the fits of the same baseline hazard function with different random-effects, as the AIC is not an appropriate measure for the model selection of random-effects[15].

Figure E1 provides the nonparametric (NP) and model-based CICs for the GG-G (left) and GG-LN (right) models for the first three gap times for the first five years, during which at least 90% of the asthma exacerbations for each gap time occurred. The GG-G model provided an excellent fit for the first gap time, as the model-based CIC (red) aligned closely with the NP CIC (black). On the other hand, the GG-LN showed a poor fit for the first gap time, as the central region of the model-based CIC and the 95% CI of the NP CIC failed to overlap for much of the range illustrated. For the subsequent gap times, both models showed deteriorating fits after the first year, most likely due to the relatively small number of follow-up events. Given that the majority of the follow-up asthma exacerbations occurred in the first gap time, we judged the goodness-of-fit of the GG-G model to be adequate for our purposes.

**Figure E1**: Nonparametric (NP) cumulative incidence curve (black) and model-based cumulative incidence curves (red) for the first three gap times for the generalised gamma model with gamma random-effects (left) and with log-normal random-effects (right). The grey shading is the 95% CI of the NP cumulative incidence curve, whereas the pink shading is the central region of the patient-specific model-based cumulative incidence curves (obtained by a functional boxplot[16]).

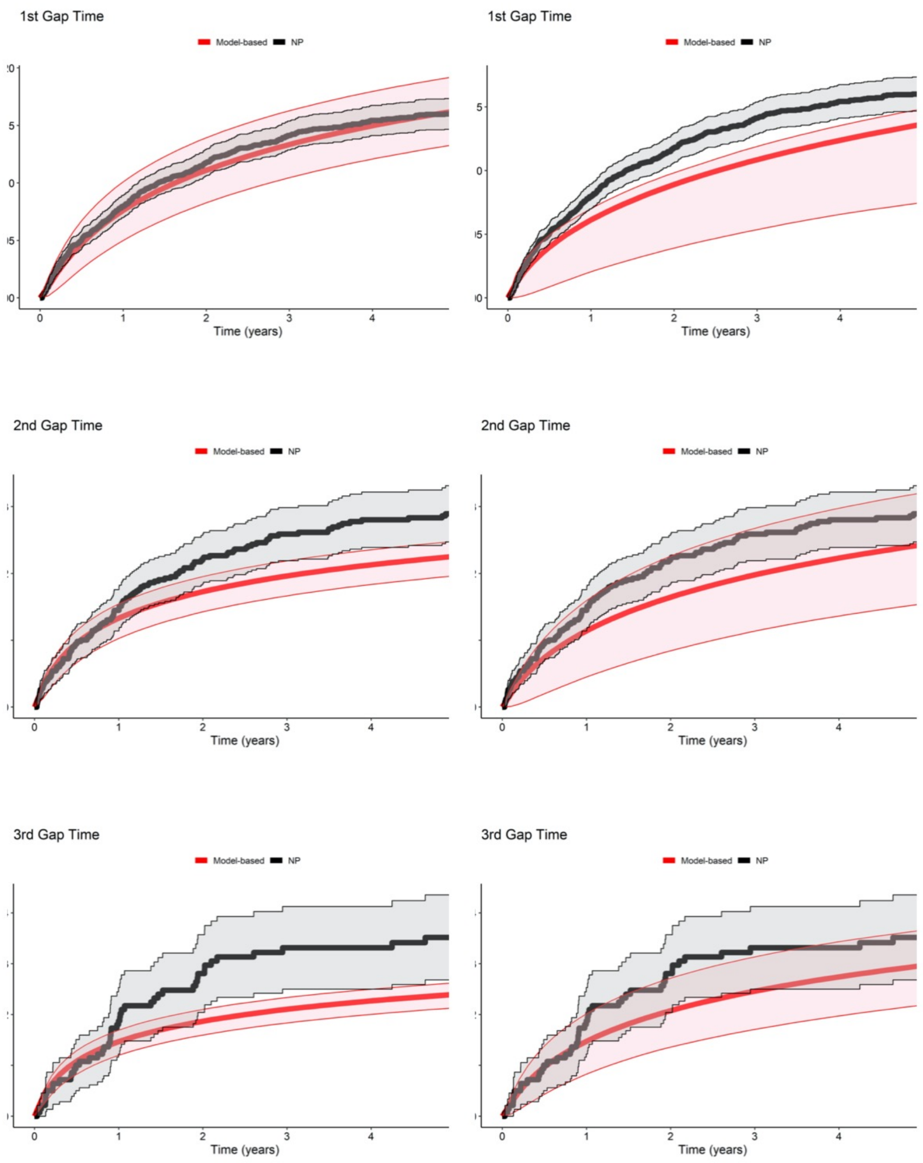

#### 2.2 Adult Group

For the adult group (n=5,459), 15% had at least one severe asthma exacerbation, 5% had two or more severe asthma exacerbations, and 2% had 3 or more severe exacerbations after their index date. About 60% of the follow-up exacerbations were first exacerbations. Unlike the paediatric group, the adult group had a significant proportion of patients who experienced severe COPD exacerbations (13%) or died (10%), requiring the modeling of competing events, as described previously. We used the exponential distribution for the baseline hazard function of both competing risks, so as to satisfy the assumption of the memoryless property of the residual times (Section 1.1). The GG/EXP/EXP-G and -LN (for asthma, COPD, and death, respectively, and with G and LN random-effects, respectively) models were fitted successfully. To compare the goodness-of-fit of the GG/EXP/EXP models with different random-effects, we relied on CICs (Figures E2 and E3).

Figure E2 provides the NP and model-based CICs of the recurrent event process of severe asthma exacerbations. For the first gap time, the plot indicates the model with G random-effects provided a better overall fit. Its model-based CIC aligned quite closely with the NP CIC for the first three years and then rose slightly above the NP CIC but generally remained within the 95% CI of the NP CIC. On the other hand, the model-based CIC for LN random-effects aligned closely with the NP CIC only for the first year and then rose above the NP CIC and outside of the 95% CI of the NP CIC. For the second and third gap times, the plots were quite comparable although, in both cases, the model with LN random-effects appeared to provide a marginally better overall fit. In summary, the differences between the two model fits are subtle. Given that the majority of the follow-up events occurred in the first gap time, we chose the model with G random-effects as our final model for the adult group, as it provided a better overall fit for the first gap time.

**Figure E2**: Nonparametric (NP) cumulative incidence curve (black) and model-based cumulative incidence curves (red) of the recurrent event process for the GG/EXP/EXP-G model (left) and the GG/EXP/EXP-LN model (right). The grey shading is the 95% CI of the NP cumulative incidence curve, whereas the pink shading is the central region of all the patient-specific model-based cumulative incidence curves (obtained by a functional boxplot[16]).

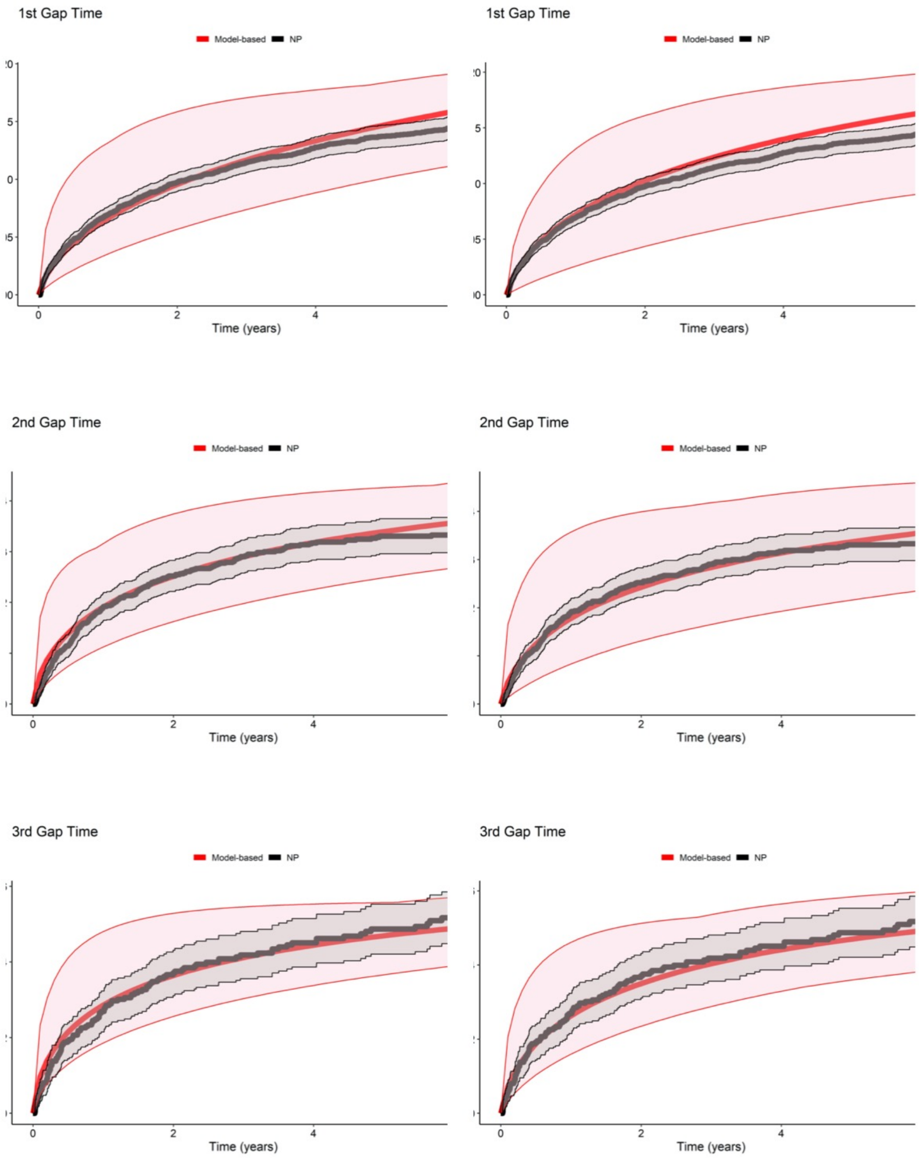

We also examined the fit of the chosen model to the terminal event data (Figure E3). For severe COPD exacerbations, the model-based CICs for the first three gap times generally remained within the 95% CI of the corresponding NP CICs. For death, the model-based CIC for the first gap time was on or slightly above the upper limit of the 95% CI of the NP CIC except for the very initial period. For the subsequent gap times, the model-based CICs remained within the 95% CI of the corresponding NP CICs. Given that our main objective was to quantify the effect of occurrence of severe asthma exacerbations on subsequent events, overall, the chosen model appeared to provide an adequate fit for the terminal event data.

**Figure E3**: Nonparametric (NP) cumulative incidence curve (black) and model-based cumulative incidence curves (red) of the terminal event processes of severe COPD exacerbations (left) and death (right) for the GG/EXP/EXP-G model. The grey shading is the 95% CI of the NP cumulative incidence curve, whereas the pink shading is the central region of the patient-specific model-based cumulative incidence curves (obtained by a functional boxplot[16]).

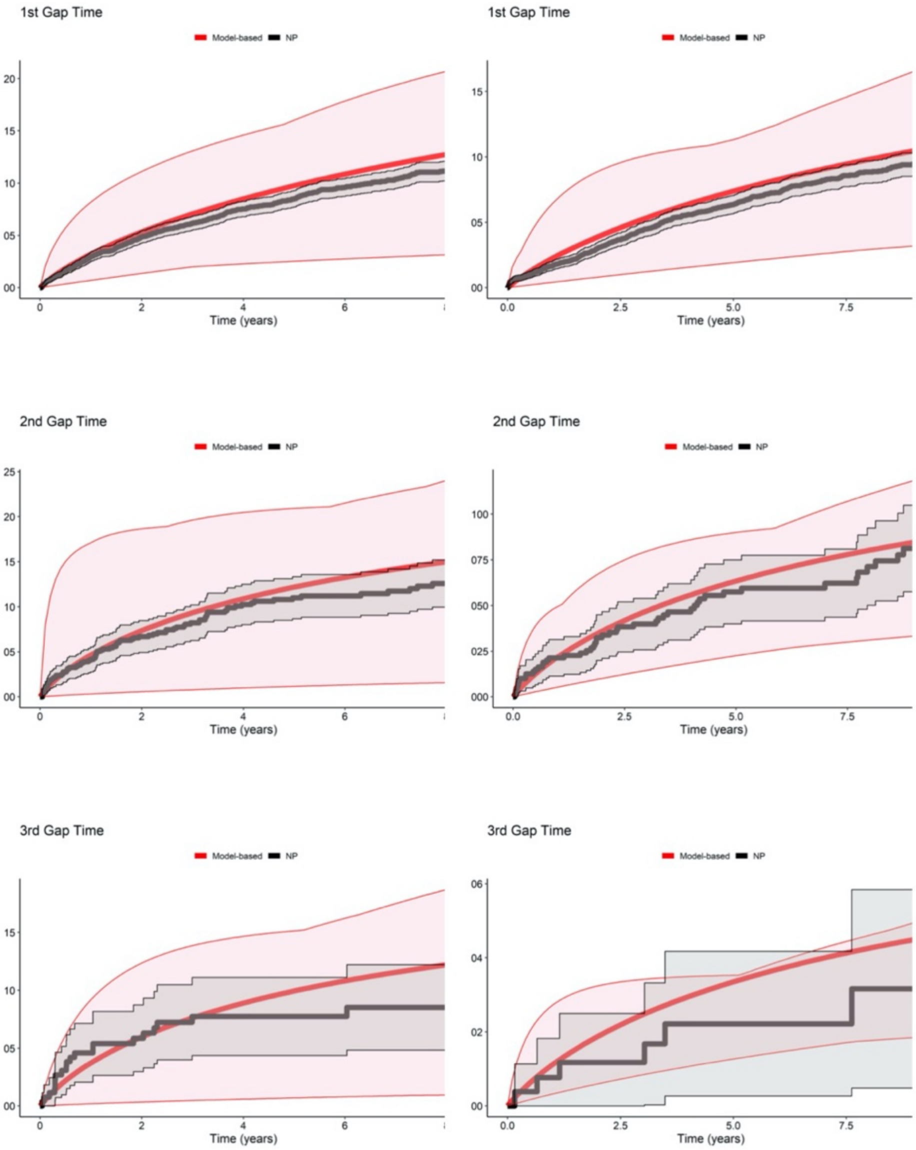

#### 2.3 Regression coefficients of the final models

Table E1 provides the estimated regression coefficients, expressed as rate acceleration factors (RAF), and parameters of the GG-G model for the paediatric group and the GG/EXP/EXP-G model for the adult group. We briefly describe the results for the asthma exacerbation events for both age groups. Biological sex was not associated with a significant change in the rate of severe asthma exacerbations for either group. On the other hand, the severity of the index asthma exacerbation, as described by the two binary covariates (length of stay and ICU admission for the index asthma exacerbation), was associated with a significant increase in the rate of subsequent events for both groups. Compared to high SES (3rd, 4th, and 5th quintiles), low SES (1st and 2nd quintiles) was not associated with an increase in the rate for the paediatric group but was associated with an increase in the rate for the adult group. Compared to the lowest index age group, the higher index age groups were associated with a decrease in the rate for both groups.

**Table E1**: Estimated rate acceleration factors (RAF) and parameters of the final models.

| | **Paediatric Group** | **Adult Group** | | |
|---|---|---|---|---|
| **Type of events** | Severe asthma exacerbation | Severe asthma exacerbation | Severe COPD exacerbation | Death |
| **Baseline hazard function** | GG | GG | EXP | EXP |
| **Time independent covariates** | **RAF (95% CI)** | | | |
| **Length of stay for the index asthma exacerbation : > 7 days** | 8.71 (1.50, 50.45)* | 1.72 (1.26, 2.36)* | - | - |
| **Charlson comorbidity index: > 0 (reference: 0)** | - | 1.40 (0.90, 2.17) | 1.91 (1.46, 2.49)* | 3.38 (2.64, 4.32)* |
| **Female** | 1.27 (0.89, 1.81) | 1.21 (0.96, 1.53) | 0.89 (0.74, 1.07) | 0.94 (0.77, 1.15) |
| **Admitted to ICU** | 0.82 (0.33, 2.00) | 2.48 (1.71, 3.58)* | - | - |
| **Index year (since 2001)** | 1.03 (0.99, 1.07) | 0.99 (0.96, 1.02) | 0.97 (0.95, 0.99)* | 0.94 (0.92, 0.97)* |
| **Social economic status: low (reference: high)** | 1.17 (0.82, 1.66) | 1.43 (1.12, 1.82)* | 1.25 (1.04, 1.51)* | 1.14 (0.93, 1.39) |
| **Social economic status: unknown (reference: high)** | - | 2.00 (1.03, 3.91)* | 1.61 (0.97, 2.69) | 1.49 (0.87, 2.55) |
| **Index age (years): 5-9 (reference: <5 )** | 0.33 (0.22, 0.51)* | - | - | - |
| **Index age (years): > 9 (reference: <5)** | 0.35 (0.21, 0.60)* | - | - | - |
| **Index age (years): 35 - 49 (reference: <35)** | - | 0.74 (0.56, 0.97)* | 23.51 (10.27, 53.82)* | 1.91 (1.24, 2.92)* |
| **Index age (years): 50 - 64 (reference: <35)** | - | 0.64 (0.48, 0.86)* | 42.05 (18.44, 95.87)* | 3.54 (2.38, 5.29)* |
| **Index age (years): > 65 (reference: <35)** | - | 0.64 (0.46, 0.88)* | 112.24 (49.26, 255.70)* | 14.75 (10.09, 21.55)* |

| Time dependent covariates | RAF (95% CI) | | | |
|---|---|---|---|---|
| **1st asthma exacerbation** | 1.80 (1.15, 2.82)* | 2.88 (1.86, 4.45)* | 1.11 (0.84, 1.46) | 0.82 (0.59, 1.14) |
| **2nd asthma exacerbation** | 0.96 (0.43, 2.17) | 1.37 (0.83, 2.26) | 0.72 (0.42, 1.23) | 0.50 (0.22, 1.14) |
| **3rd+ asthma exacerbation** | 0.93 (0.29, 3.00) | 0.87 (0.49, 1.55) | 0.70 (0.36, 1.37) | 0.89 (0.31, 2.54) |
| **Model parameters** | **Estimate (95% CI)** | | | |
| **scale** | 3.74 (3.05, 4.42) | 2.20 (1.90, 2.49) | - | - |
| **shape** | -3.47 (-4.58, -2.37) | 0.45 (0.24, 0.66) | - | - |
| **theta** | 0.68 (0.45, 1.04) | 0.86 (0.67, 1.10) | | |
| **COPD association factor** | - | 0.78 (0.57, 0.98) | | |
| **Death association factor** | - | 0.61 (0.36, 0.86) | | |

* significant at 0.05 level; COPD: chronic obstructive pulmonary disease; GG: generalised gamma; EXP: exponential; RAF: rate acceleration factors; ICU: intensive care unit; CI: confidence interval

### 3 Sensitivity analysis

For the adult group, we assumed the memoryless property of the residual times for the terminal event processes, restricting the choice of their baseline hazard functions to the exponential distribution[6]. To check on the impact of that restriction, we ran a sensitivity analysis where we used calendar times for both terminal events, thereby allowing more flexible modeling of those events. For the sake of simplicity, we excluded the time dependent covariates indicating the number of the follow-up asthma exacerbations from the hazard functions for the two terminal events.

Let $V_i$ be the time since the index date and $V_i^*$ be the time with asthma exacerbation durations since the index date excluded. Let $FU_i^* = FU_i - \sum_{j=1}^{m_i} B_{i,j}$ be the follow-up time with asthma exacerbation durations excluded. Following the notation used in Section 1, the AFT-type joint frailty model with calendar times for the terminal events adapted from [4] is

$$\begin{cases} h\left(w_{i,j}^*|\epsilon_i\right) = h_0\left(w_{i,j}^* \, \epsilon_\mathrm{i} \exp\left(\beta_A^T Z_{i,j}^A\right)\right) \epsilon_\mathrm{i} \exp\left(\beta_A^T Z_{i,j}^A\right) & \textbf{\textit{asthma}} \\ h^C\left(v_i^*|\epsilon_i\right) = h_0^C\left(v_i^* \epsilon_\mathrm{i}^{\alpha_C} \exp(\beta_C^T Z_{i,j}^C)\right) \epsilon_\mathrm{i}^{\alpha_C} \exp\left(\beta_C^T Z_{i,j}^C\right) & \textbf{\textit{COPD}} \\ h^D\left(v_i|\epsilon_i\right) = h_0^D\left(v_i \epsilon_\mathrm{i}^{\alpha_D} \exp(\beta_D^T Z_{i,j}^D)\right) \epsilon_\mathrm{i}^{\alpha_D} \exp\left(\beta_D^T Z_{i,j}^D\right) & \textbf{\textit{death}} \end{cases}$$

We derive the conditional likelihood for patient $i$ given the random-effect $\epsilon_i$. Using the calendar times for the terminal events implies that the contribution to the likelihood is simply $f_i\left(w_{i,j}^*\right)$ for experiencing the $j$th recurrent event. For the last $(m_i + 1)$th event, the patient either experiences a severe COPD exacerbation, dies, or exits the study (censored) without experiencing another severe asthma exacerbation. Thus, the conditional likelihood for patient $i$ is

$$
\begin{aligned}
L_i(\Phi|\epsilon_i) = &\left[\, \Pi_{j=1}^{m_i} f_i\left(w_{i,j}^*\right)\right] \\
&\times \left[\, S_i\left(w_{i,m_i+1}^*\right)\right. \\
&\times \left(f_i^C(FU_i^*)\right)^{\delta_i^C} \left(S_i^C(FU_i^*)\right)^{1-\delta_i^C} \\
&\times \left(f_i^D(FU_i)\right)^{\delta_I^D} \left(S_i^D(FU_i)\right)^{1-\delta_i^D} \left.\right].
\end{aligned}
$$

The marginal likelihood for the whole sample can be obtained following the same steps described in Section 1.

We fitted the joint frailty model with calendar times for the terminal events. We used the same baseline hazard functions and random-effect distribution from the final model of the adult group, as well as the more flexible generalised gamma baseline hazard functions for the terminal event processes. The Akaike information criterion comparing the GG/EXP/EXP-G and GG/GG/GG-G models indicated that the more flexible baseline hazard functions provided a better fit to the data, so we chose GG/GG/GG-G as the sensitivity model to compare with the earlier final model for the adult group.

Table E2 provides the estimated regression coefficients (expressed as RAFs) and parameters of the sensitivity model using the calendar times for the terminal events. The estimated RAFs of the time independent covariates for all three processes were very close to those of the final model (Table E1). Figure E4 provides the estimated RAFs of the time dependent covariates indicating the number of the follow-up asthma exacerbation for the recurrent event process in the final and sensitivity models. For each follow-up asthma exacerbation, the estimated RAF was somewhat lower in the sensitivity model, but our conclusions remain unchanged: the occurrence of the first follow-up asthma exacerbation is associated with a significant increase in the rate of future events, and the effects of the subsequent follow-up exacerbations are not statistically significant. In conclusion, the sensitivity analysis suggests that our conclusions are not overly sensitive to the model formulation used for the terminal event processes.

**Table E2**: Estimated rate acceleration factors (RAF) and parameters of the sensitivity model.

| **Type of events** | Severe asthma exacerbation | Severe COPD exacerbation | Death |
|---|---|---|---|
| **Baseline hazard function** | GG | GG | GG |
| **Time independent covariates** | **RAF (95% CI)** | | |
| **Length of stay for the index asthma exacerbation : > 7 days** | 1.88 (1.33, 2.64)* | - | - |
| **Charlson comorbidity index: > 0 (reference: 0)** | 1.36 (0.85, 2.18) | 1.93 (1.38, 2.71)* | 3.52 (2.56, 4.83)* |
| **Female** | 1.22 (0.95, 1.57) | 0.88 (0.70, 1.10) | 0.92 (0.74, 1.15) |
| **Admitted to ICU** | 2.82 (1.89, 4.20)* | - | - |
| **Index year (since 2001)** | 0.99 (0.96, 1.02) | 0.96 (0.93, 0.98)* | 0.94 (0.91, 0.97)* |
| **Social economic status: low (reference: high)** | 1.50 (1.16, 1.94)* | 1.30 (1.04, 1.64)* | 1.11 (0.89, 1.38) |
| **Social economic status: unknown (reference: high)** | 2.12 (1.03, 4.39)* | 1.73 (0.92, 3.25) | 1.43 (0.79, 2.58) |
| **Index age (years): 35 - 49 (reference: <35)** | 0.71 (0.53, 0.94)* | 34.43 (13.60, 87.13)* | 2.00 (1.21, 3.32)* |
| **Index age (years): 50 - 64 (reference: <35)** | 0.61 (0.44, 0.83)* | 63.29 (25.10, 159.59)* | 3.68 (2.27, 5.95)* |
| **Index age (years): > 65 (reference: <35)** | 0.53 (0.38, 0.74)* | 179.25 (70.36, 456.69)* | 16.43 (10.28, 26.25)* |
| **Time dependent covariates** | **RAF (95% CI)** | | |
| **1st asthma exacerbation** | 1.87 (1.07, 3.24)* | - | - |
| **2nd asthma exacerbation** | 1.01 (0.58, 1.75) | - | - |
| **3rd+ asthma exacerbation** | 0.64 (0.37, 1.09) | - | - |
| **Model parameters** | **Estimate (95% CI)** | | |
| **scale** | 2.08 (1.71, 2.45) | 1.66 (1.37, 1.95) | 1.16 (0.84, 1.49) |
| **shape** | 0.50 (0.19, 0.81) | 0.58 (0.33, 0.82) | 0.98 (0.61, 1.36) |
| **theta** | 1.00 (0.83, 1.30) | | |
| **COPD association factor $\alpha_C$** | 0.54 (0.31, 0.77) | | |
| **Death association factor $\alpha_D$** | 0.07 (-0.13, 0.28) | | |

* significant at 0.05 level; GG: generalised gamma; RAF: rate acceleration factors; ICU: intensive care unit; CI: confidence interval

**Figure E4**: Estimated rate acceleration factors for severe asthma exacerbations in the final (orange) and sensitivity (blue) models for the adult group.

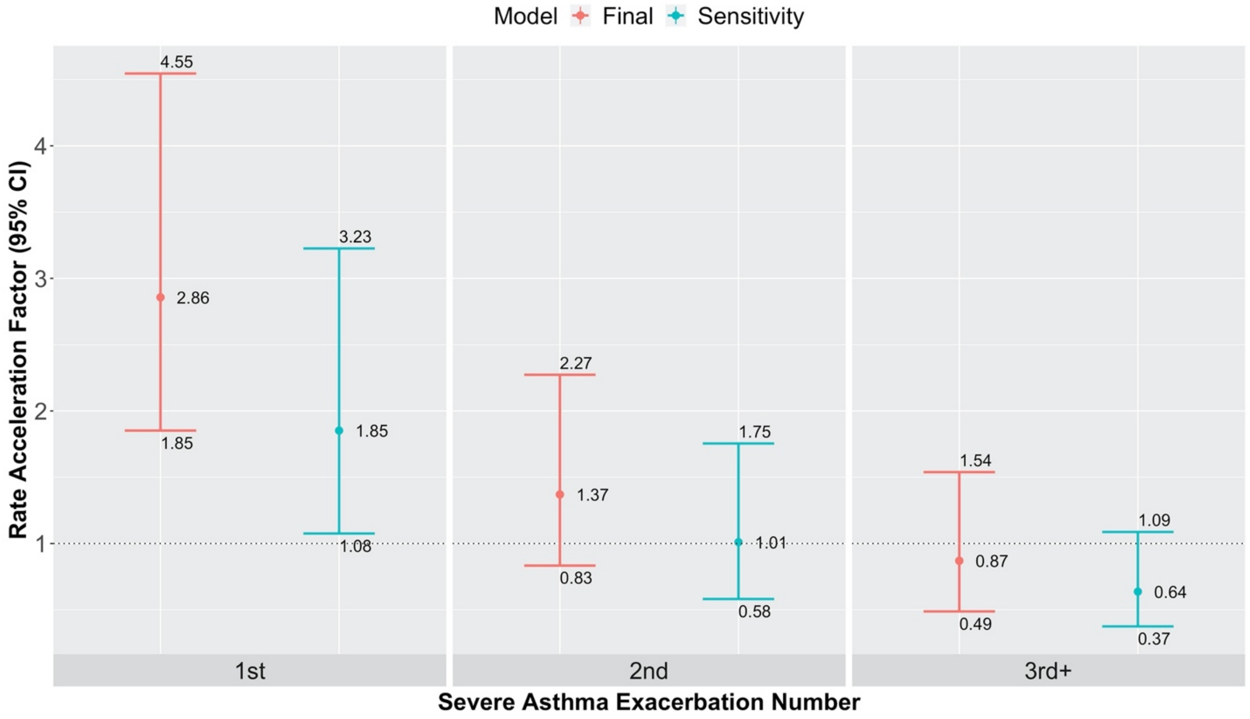


CI: confidence interval

### 4 SAS codes

We provide the SAS codes for fitting the GG/EXP/EXP-G, GG/EXP/EXP-LN, and GG/GG/GG-G models. For initialization, we used a grid of values around the maximum likelihood estimates obtained from fitting each hazard function separately without random-effects. We transformed the variance parameter, $\theta > 0$, to the log scale so that the transformed variance parameter is unbounded. We recommend transforming the scale parameter as well if its estimate lies near the boundary value of 0, so as to avoid convergence issues.

#### GG/EXP/EXP-G model

```
TITLE "joint frailty, gg, exp, exp, G";
PROC NLMIXED DATA=df_filtered gCONV=0 QPOINTS=5 TECHNIQUE=NRRIDG
MAXTIME=10000 MAXFUNC=10000 MAXIT=10000 GCONV=1E-8 NTHREADS=7;
	/* set initial parameter values*/

			p = cdf("NORMAL",z);
			if p > 0.99999 then p=0.99999;
			g2 = quantile('GAMMA',p,exp(inv_ln_theta));
			g = g2 *1/exp(inv_ln_theta);

	tmp_A = b0_A+
				age1_A*age1+ age2_A*age2+ age3_A*age3+
				cohort_year_A*cohort_year+
				SES_low_A*SES_low+ SES_unk_A*SES_unk+
				CCI_A*CCI+
				asthma_1_A*asthma_1+ asthma_2_A*asthma_2+
asthma_3_A*asthma_3+
				baseline_duration_A * baseline_duration+
				ICU_A * ICU +
				female_A*female;

			lin_A = -tmp_A + log(g);

			if (shape_A>=0) then
		l_S_A=log(1-
cdf('gamma',gaptime_asthma**(shape_A/scale_A),1/(shape_A*shape_A),shape_A*shape_A*ex
p(lin_A*shape_A/scale_A)));
	if (shape_A<0) then

l_S_A=log(cdf('gamma',gaptime_asthma**(shape_A/scale_A),1/(shape_A*shape_A),shape_A*s
hape_A*exp(lin_A*shape_A/scale_A)));
```

```
		l_f_A = log(abs(shape_A))-log(scale_A)-lgamma((shape_A**(-
2)))+(shape_A**(-2))*((-2)*log(abs(shape_A))+(shape_A/scale_A)*(-
lin_A+log(gaptime_asthma)))-(shape_A**(-2))*((exp(-
lin_A)*gaptime_asthma)**(shape_A/scale_A));


		tmp_C = b0_C+
			age1_C*age1+ age2_C*age2+ age3_C*age3+
			cohort_year_C*cohort_year+
			SES_low_C*SES_low+ SES_unk_C*SES_unk+
			CCI_C*CCI+
			asthma_1_C*asthma_1+ asthma_2_C*asthma_2+
asthma_3_C*asthma_3+
			female_C*female;

		lin_C = -tmp_C + c*log(g);

		alpha_C = exp(-lin_C);

		l_S_C=-alpha_C*gaptime_copd;

		l_f_C= log(alpha_C) +l_S_C;

		tmp_D = b0_D+
			age1_D*age1+ age2_D*age2+ age3_D*age3+
			cohort_year_D*cohort_year+
			SES_low_D*SES_low+ SES_unk_D*SES_unk+
			CCI_D*CCI+
			asthma_1_D*asthma_1+ asthma_2_D*asthma_2+
asthma_3_D*asthma_3+
			female_D*female;

		lin_D = -tmp_D + d*log(g);

		alpha_D = exp(-lin_D);

		l_S_D=-alpha_D*gaptime_death;

		l_f_D= log(alpha_D) +l_S_D;

		if (type=0) then
			ll = l_S_D + l_S_C + l_S_A;
		if (type=1) then
			ll = l_S_D + l_S_C + l_f_A;
		if (type=2) then
			ll = l_S_D + l_f_C + l_S_A;
```

```
        if (type=3) then
                ll = l_f_D + l_S_C + l_S_A;

        RANDOM z ~ NORMAL(0 , 1) SUBJECT=STUDYID OUT=_Z;

        MODEL gaptime_asthma ~ general(ll);

        PREDICT lin_A OUT=_lin_A;
        PREDICT lin_C OUT=_lin_C;
        PREDICT lin_D OUT=_lin_D;

        ESTIMATE 'theta' 1/exp(inv_ln_theta);

    ODS OUTPUT ParameterEstimates=_tmp1;
    ODS OUTPUT AdditionalEstimates=_tmp2;
        ODS OUTPUT FitStatistics=_fit;


RUN;
ods rtf close;
```

#### GG/EXP/EXP-LN model

```
TITLE "joint frailty, gg, exp, exp, LN";
PROC NLMIXED DATA=df_filtered gCONV=0 QPOINTS=5 TECHNIQUE=NRRIDG
MAXTIME=10000 MAXFUNC=10000 MAXIT=10000 GCONV=1E-8 NTHREADS=7;

        /* initial parameter values*/

    tmp_A = b0_A+
                age1_A*age1+ age2_A*age2+ age3_A*age3+
                cohort_year_A*cohort_year+
                SES_low_A*SES_low+ SES_unk_A*SES_unk+
                CCI_A*CCI+
                asthma_1_A*asthma_1+ asthma_2_A*asthma_2+
asthma_3_A*asthma_3+
                baseline_duration_A * baseline_duration+
                ICU_A * ICU +
                female_A*female;

        lin_A = -tmp_A + z;

        if (shape_A>=0) then
      l_S_A=log(1-
cdf('gamma',gaptime_asthma**(shape_A/scale_A),1/(shape_A*shape_A),shape_A*shape_A*ex
p(lin_A*shape_A/scale_A)));
```

```
        if (shape_A<0) then

l_S_A=log(cdf('gamma',gaptime_asthma**(shape_A/scale_A),1/(shape_A*shape_A),shape_A*s
hape_A*exp(lin_A*shape_A/scale_A)));
                l_f_A = log(abs(shape_A))-log(scale_A)-lgamma((shape_A**(-
2)))+(shape_A**(-2))*((-2)*log(abs(shape_A))+(shape_A/scale_A)*(-
lin_A+log(gaptime_asthma)))-(shape_A**(-2))*((exp(-
lin_A)*gaptime_asthma)**(shape_A/scale_A));


                tmp_C = b0_C+
                        age1_C*age1+ age2_C*age2+ age3_C*age3+
                        cohort_year_C*cohort_year+
                        SES_low_C*SES_low+ SES_unk_C*SES_unk+
                        CCI_C*CCI+
                        asthma_1_C*asthma_1+ asthma_2_C*asthma_2+
asthma_3_C*asthma_3+
                        female_C*female;

                lin_C = -tmp_C + c*z;

                alpha_C = exp(-lin_C);

                l_S_C=-alpha_C*gaptime_copd;

                l_f_C= log(alpha_C) +l_S_C;

                tmp_D = b0_D+
                        age1_D*age1+ age2_D*age2+ age3_D*age3+
                        cohort_year_D*cohort_year+
                        SES_low_D*SES_low+ SES_unk_D*SES_unk+
                        CCI_D*CCI+
                        asthma_1_D*asthma_1+ asthma_2_D*asthma_2+
asthma_3_D*asthma_3+
                        female_D*female;

                lin_D = -tmp_D + d*z;

                alpha_D = exp(-lin_D);

                l_S_D=-alpha_D*gaptime_death;

                l_f_D= log(alpha_D) +l_S_D;


                if (type=0) then
```

```
		ll = l_S_D + l_S_C + l_S_A;
	if (type=1) then
		ll = l_S_D + l_S_C + l_f_A;
	if (type=2) then
		ll = l_S_D + l_f_C + l_S_A;
	if (type=3) then
		ll = l_f_D + l_S_C + l_S_A;

	RANDOM z ~ NORMAL(0 , exp(ln_theta)) SUBJECT=STUDYID OUT=_Z;

	MODEL gaptime_asthma ~ general(ll);

	PREDICT lin_A OUT=_lin_A;
	PREDICT lin_C OUT=_lin_C;
	PREDICT lin_D OUT=_lin_D;

	ESTIMATE 'theta' exp(ln_theta);

  ODS OUTPUT ParameterEstimates=_tmp1;
  ODS OUTPUT AdditionalEstimates=_tmp2;
	ODS OUTPUT FitStatistics=_fit;


RUN;
ods rtf close;
```

#### Sensitivity model (GG/GG/GG-G)

```
TITLE "joint frailty, gg, gg, gg, G, tte";
PROC NLMIXED DATA=df_filtered gCONV=0 QPOINTS=5 TECHNIQUE=CONGRA
MAXTIME=10000 MAXFUNC=10000 MAXIT=10000 GCONV=1E-8 NTHREADS=7;
	/* set initial parameter values */

	bounds scale_A scale_D scale_C > 0;

	p = cdf("NORMAL",z);
	if p > 0.99999 then p=0.99999;
	g2 = quantile('GAMMA',p,exp(inv_ln_theta));
	g = g2 *1/exp(inv_ln_theta);

  tmp_A = b0_A+
		age1_A*age1+ age2_A*age2+ age3_A*age3+
		cohort_year_A*cohort_year+
		SES_low_A*SES_low+ SES_unk_A*SES_unk+
		CCI_A*CCI+
```

```
                    asthma_1_A*asthma_1+ asthma_2_A*asthma_2+
asthma_3_A*asthma_3+
                    baseline_duration_A * baseline_duration+
                    ICU_A * ICU +
                    female_A*female;

                lin_A = -tmp_A + log(g);

                if (shape_A>=0) then
            l_S_A=log(1-
cdf('gamma',gaptime_asthma**(shape_A/scale_A),1/(shape_A*shape_A),shape_A*shape_A*ex
p(lin_A*shape_A/scale_A)));
        if (shape_A<0) then

l_S_A=log(cdf('gamma',gaptime_asthma**(shape_A/scale_A),1/(shape_A*shape_A),shape_A*s
hape_A*exp(lin_A*shape_A/scale_A)));
                l_f_A = log(abs(shape_A))-log(scale_A)-lgamma((shape_A**(-
2)))+(shape_A**(-2))*((-2)*log(abs(shape_A))+(shape_A/scale_A)*(-
lin_A+log(gaptime_asthma)))-(shape_A**(-2))*((exp(-
lin_A)*gaptime_asthma)**(shape_A/scale_A));

                tmp_C = b0_C+
                    age1_C*age1+ age2_C*age2+ age3_C*age3+
                    cohort_year_C*cohort_year+
                    SES_low_C*SES_low+ SES_unk_C*SES_unk+
                    CCI_C*CCI+
                    female_C*female;

                lin_C = -tmp_C + c*log(g);

                if (shape_C>=0) then
            l_S_C=log(1-
cdf('gamma',tte_copd**(shape_C/scale_C),1/(shape_C*shape_C),shape_C*shape_C*exp(lin_C*
shape_C/scale_C)));
        if (shape_C<0) then

l_S_C=log(cdf('gamma',tte_copd**(shape_C/scale_C),1/(shape_C*shape_C),shape_C*shape_C
*exp(lin_C*shape_C/scale_C)));
                l_f_C = log(abs(shape_C))-log(scale_C)-lgamma((shape_C**(-
2)))+(shape_C**(-2))*((-2)*log(abs(shape_C))+(shape_C/scale_C)*(-lin_C+log(tte_copd)))-
(shape_C**(-2))*((exp(-lin_C)*tte_copd)**(shape_C/scale_C));

                tmp_D = b0_D+
                    age1_D*age1+ age2_D*age2+ age3_D*age3+
                    cohort_year_D*cohort_year+
                    SES_low_D*SES_low+ SES_unk_D*SES_unk+
```

```
                    CCI_D*CCI+
                    female_D*female;

            lin_D = -tmp_D + d*log(g);

            if (shape_D>=0) then
        l_S_D=log(1-
cdf('gamma',tte_death**(shape_D/scale_D),1/(shape_D*shape_D),shape_D*shape_D*exp(lin_D
*shape_D/scale_D)));
    if (shape_D<0) then

l_S_D=log(cdf('gamma',tte_death**(shape_D/scale_D),1/(shape_D*shape_D),shape_D*shape_
D*exp(lin_D*shape_D/scale_D)));
            l_f_D = log(abs(shape_D))-log(scale_D)-lgamma((shape_D**(-
2)))+(shape_D**(-2))*((-2)*log(abs(shape_D))+(shape_D/scale_D)*(-lin_D+log(tte_death)))-
(shape_D**(-2))*((exp(-lin_D)*tte_death)**(shape_D/scale_D));

            if (type=0) then
                    ll = l_S_D + l_S_C + l_S_A;
            if (type=1) then
                    ll = l_f_A;
            if (type=2) then
                    ll = l_S_D + l_f_C + l_S_A;
            if (type=3) then
                    ll = l_f_D + l_S_C + l_S_A;

            RANDOM z ~ NORMAL(0 , 1) SUBJECT=STUDYID OUT=_Z;

            MODEL gaptime_asthma ~ general(ll);

            PREDICT lin_A OUT=_lin_A;
            PREDICT lin_C OUT=_lin_C;
            PREDICT lin_D OUT=_lin_D;

            ESTIMATE 'theta' 1/exp(inv_ln_theta);

    ODS OUTPUT ParameterEstimates=_tmp1;
    ODS OUTPUT AdditionalEstimates=_tmp2;
            ODS OUTPUT FitStatistics=_fit;


RUN;
ods rtf close;
```

#### References


1. Silva GE, Sherrill DL, Guerra S, Barbee RA. Asthma as a risk factor for COPD in a longitudinal study. *Chest* Elsevier; 2004; 126: 59–65.

2. Diaz-Guzman E, Khosravi M, Mannino DM. Asthma, chronic obstructive pulmonary disease, and mortality in the US population. *COPD: Journal of Chronic Obstructive Pulmonary Disease* Taylor & Francis; 2011; 8: 400–407.

3. Schuster NA, Hoogendijk EO, Kok AAL, Twisk JWR, Heymans MW. Ignoring competing events in the analysis of survival data may lead to biased results: a nonmathematical illustration of competing risk analysis. *Journal of Clinical Epidemiology* 2020; 122: 42–48.

4. Huang X, Liu L. A joint frailty model for survival and gap times between recurrent events. *Biometrics* 2007; 63: 389–397.

5. Wei L-J. The accelerated failure time model: a useful alternative to the Cox regression model in survival analysis. *Statistics in Medicine* Wiley Online Library; 1992; 11: 1871–1879.

6. Nelsen RB. Consequences of the memoryless property for random variables. *The American Mathematical Monthly* Mathematical Association of America; 1987; 94: 981–984.

7. SAS Institute Inc. SAS/STAT 14.1 User's Guide. Cary, NC; 2015.

8. Lu L, Liu C. Analysis of correlated recurrent and terminal events data in SAS. *Proceedings of Northeastern SAS Users' Group 2008 Conference* 2008.

9. Liu L, Huang X. The use of Gaussian quadrature for estimation in frailty proportional hazards models. *Statistics in Medicine* 2008; 27: 2665–2683.

10. Sadatsafavi M, Xie H, Etminan M, Johnson K, FitzGerald JM, Canadian Respiratory Research Network. The association between previous and future severe exacerbations of chronic obstructive pulmonary disease: updating the literature using robust statistical methodology. *PloS One* 2018; 13: e0191243.

11. Angus JE. The probability integral transform and related results. *SIAM review* SIAM; 1994; 36: 652–654.

12. Nelson KP, Lipsitz SR, Fitzmaurice GM, Ibrahim J, Parzen M, Strawderman R. Use of the probability integral transformation to fit nonlinear mixed-effects models with nonnormal random effects. *Journal of Computational and Graphical Statistics* Taylor & Francis; 2006; 15: 39–57.

13. Cox C, Chu H, Schneider MF, Munoz A. Parametric survival analysis and taxonomy of hazard functions for the generalized gamma distribution. *Statistics in Medicine* Wiley Online Library; 2007; 26: 4352–4374.

14. Kleinbaum DG, Klein M. Survival Analysis. 3rd ed. Springer;

15. Greven S, Kneib T. On the behaviour of marginal and conditional AIC in linear mixed models. *Biometrika* Oxford Academic; 2010; 97: 773–789.

16. Sun Y, Genton MG. Functional boxplots. *Journal of Computational and Graphical Statistics* Taylor & Francis; 2011; 20: 316–334.
